# Neck pain, dry eye and Sjögren’s syndrome in Latin American students during the first wave of COVID-19: Frequencies and associated factors

**DOI:** 10.1101/2023.03.21.23287540

**Authors:** Christian R. Mejia, Briggitte Gutarra-Laureano, Ingrid L. Zorrilla-Lizana, Dennis Arias-Chavez, Maria F. Fernandez, Claudia A. Vera, Martin A. Vilela-Estrada, Victor Serna-Alarcón, Tatiana Requena, Lysien Ivania Zambrano, Eleonora Espinoza Turcios

**Affiliations:** Universidad Norbert Wiener. Translational Medicine Research Centre. Lima, Peru; Universidad Continental. Huancayo, Peru; Universidad Privada de Tacna. Tacna, Peru; Instituto Nacional de Oftalmología. Lima, Peru; Escuela Profesional de Medicina Humana. Universidad Privada Antenor Orrego. Trujillo, Peru; Hospital Regional José Cayetano Heredia, EsSalud. Piura, Peru; Universidad Cesar Vallejo. Trujillo, Peru; Institute for Research in Medical Sciences and Right to Health (ICIMEDES)/Scientific Research Unit (UIC), Faculty of Medical Sciences (FCM), National Autonomous University of Honduras (UNAH). Tegucigalpa, Honduras

**Author notes:** **Corresponding authors:** Eleonora Espinoza Turcios, Instituto de Investigación en Ciencias Médicas y Derecho a la Salud (ICIMEDES)/Unidad de Investigación Científica (UIC), Edificio CM1, FCM, Calle La Salud, contiguo al Hospital Escuela, Tegucigalpa, M. D.C., Honduras, C.A. Código Postal: 11101.

**Keywords:** Education, Neck Pain, Dry Eye, Sjögren’s Syndrome, Students

## Abstract

**Introduction:** Virtual classes brought many changes to the lives of students, not only the fact of being more exposed to screens, but also because of the repercussions.

**Aim:** To determine the factors associated with suffering from neck pain, dry eye and Sjögren’s syndrome in students in Latin America during the first wave of COVID-19.

**Methodology:** Analytical cross-sectional study, using the COM and DEQ-5 scales, neck pain and dry eye/Sjögren’s syndrome, respectively, were measured; socio-educational variables were associated with them.

**Discussion:** Of the 3939 students, those who lived in Panama, Chile and Bolivia were the ones who suffered the most from these pathologies. These pathologies were associated with the greater number of hours of computer use (all values p<0,001) and sex (all values p<0,002), medical students had more frequent dry eye and Sjögren’s syndrome (both p<0,031), Graduate students had more neck pain (p<0.001), but college students had less dry eye (p=0.025) and those at private universities had more neck pain (p=0.024).

**Discussion:** Important results of these three pathologies were found, this serves so that students can be evaluated in depth in each university, for a specialized diagnosis and try to avoid medium and long-term consequences for the constant use of electronic devices.

**Conclusion:** Neck pain, dry eye and Sjögren’s syndrome in students were associated with more hours of computer use and female sex, medical students had more frequent dry eye and Sjögren’s syndrome, graduate students had more neck pain, university students had less dry eye and those from private universities had more neck pain.

## Introduction

The pandemic generated a sudden change in education worldwide, according to UNESCO, the crisis affected about 363 million students worldwide, including 57.8 million students in higher education, in addition, a large percentage of these moved away from the classroom (1). All due to the quarantine, curfews and other social restrictions, which prevented him from going to the classrooms in person (2). In all this context, it was necessary to give an unexpected impulse to new forms of teaching. (3). Although, prior to the pandemic, some universities, schools, undergraduate and graduate programs had already opted for online learning, this never happened on such a large scale. (4, 5).

Therefore, educational institutions had to adapt quickly and, although at first, some institutions found it difficult to enter the world of online learning, after a while almost all had to resort to platforms and other virtual means for the dictation of their classes; thus opting for a comprehensive system of digital education (6, 7). This new model of education reduced the learning loss, but also generated some problems, including distraction, poor internet signal, equipment not suitable for receiving virtual classes, disinterest on the part of students, the large amount of plagiarism / copy or other bad practices in exams, among many others; which generated an incomplete learning process (8, 9).

As a result of this new form of teaching-learning, physio-ergonomic problems have been reported, due to the use of study environments that were not prepared or correctly set to receive classes virtually (10, 11). Since, the students had to have an appropriate computer, screen filters, ergonomic mice, seats that can be graduated according to the physiognomy of the student, an optimal lighting system, pauses / breaks from time to time; those that together avoid or reduce health problems (11, 12). Although there were not many reports in students, previous problems had already been reported in workplaces, such as body aches, backaches, dry eyes, low back pain, carpal tunnel syndrome; among those who were for a long time in front of screens (12, 13).

For example, neck pain was reported between 10.4-21.3%, with a higher incidence in office and computer workers, neck pain between 0.4-86.8% (mean: 23.1%); being higher in women, high-income countries and urban areas (14). This situation has been little studied among university students, who were exposed the first 6 months of the pandemic to this radical change, especially in a population as diverse and varied as the Latin American region (15, 16). Therefore, the objective of the research was to determine the factors associated with suffering from neck pain, dry eye and Sjögren’s syndrome in Latin American students during the first wave of COVID-19.

## Methodology

Analytical cross-sectional research was conducted. The surveyed population were students from the countries of Peru, Chile, Paraguay, Mexico, Bolivia, Panama, Ecuador, Costa Rica, El Salvador, and Honduras; These were recruited during the months of June, July and August of the year 2020. Participants who agreed to be part of the research, who said they lived in some of these countries at the precise time the respondent was conducted, were included. Fewer than 300 students were excluded because they did not provide answers for the main tests (for the assessment of neck pain, dry eye or Sjögren’s syndrome).

Non-random sampling was used. The statistical power of the association of each of the factors was obtained, the power was not enough for neck pain versus having technical studies (3%), or university (8%), or for studying medicine (76%); likewise, an adequate power was not obtained for the crossing of having dry eye or Sjögren’s syndrome versus having university studies (67% and 64%, respectively); Therefore, these unique crossovers should be taken with caution for the discussion of results. The information was obtained, a quality control of the data was carried out and statistical analysis was carried out. All this was done in a sheet of the Microsoft Excel program, then the information was exported to a sheet of the Stata version 16 program (licensed by the group’s statistician).

### Test

Non-random sampling was used. The statistical power of the association of each of the factors was obtained, the power was not enough for neck pain versus having technical studies (3%), or university (8%), or for studying medicine (76%); likewise, an adequate power was not obtained for the crossing of having dry eye or Sjögren’s syndrome versus having university studies (67% and 64%, respectively); Therefore, these unique crossovers should be taken with caution for the discussion of results. The information was obtained, a quality control of the data was carried out and statistical analysis was carried out. All this was done in a sheet of the Microsoft Excel program, then the information was exported to a sheet of the Stata version 16 program (licensed by the group’s statistician) (17, 18).

For the results of dry eye and Sjögren’s syndrome, the DEQ-5 questionnaire was used, where through 5 questions it was possible to evaluate the suffering of these two pathologies, through this questionnaire that was validated in the Mexican population and in other places (19, 20). To obtain dry eye, one had to have a score greater than 6 and to determine that one had Sjögren’s syndrome a score greater than 12; This as a result of the total sum of each of the questions (21).

In addition, the variables of the country of residence, the sex of the respondents, the age, the degree of maximum education with which they had or were studying, the type of university in which they were enrolled, the number of years of study they had to date, if they studied human medicine, the number of hours they used the cell phone and the computer per day were counted. All these variables first went through a descriptive statistical analysis, where the frequency of presentation of each of the three pathologies according to the countries was obtained, with this it was possible to build a first figure. Then we proceeded to construct the tables of the crosses, where the crossing versus the socio-educational variables was obtained for each of the pathologies; This is where the frequency and percentage for the categorical variables were obtained, as well as the median and the interquartile range for the independent quantitative variables (this due to their condition of non-normality, evaluated through the Shapiro-Wilk statistical test).

Bivariate and multivariate analyses were also obtained for the crossing of each pathology according to the variables, this generated with the use of generalized linear models, where the Poisson family was used, the log link function, the adjustment for robust variants and by the country of residence; with this, the prevalence ratios were obtained (crude for the bivariate models and adjusted for the multivariate model), their 95% confidence intervals (95% CI) and their P values; for a variable to move from the bivariate to the multivariate model, it had to have a p<0.05 value. At all times the significance value of 0.05 was considered and we always worked with a confidence level of 95%.

### Ethics

Ethics were always respected, surveys were anonymous, and participants were free to participate or not participate in the research. The research was approved by the Bioethics Committee of the Antenor Orrego Private University, with Resolution No. 0013-2022-UPAO.

## Results

Of the 3939 students surveyed in some Latin American countries, the countries that had the highest percentages of neck pain were Panama, Chile, and Peru (with ranges 20-40%), for dry eye were Chile, Bolivia, and Costa Rica (with ranges 29-65%) and for dry eye, Sjögren’s syndrome were Chile, Bolivia and Panama (with ranges 16-35%). **Fig 1**.

**Figure 1.**
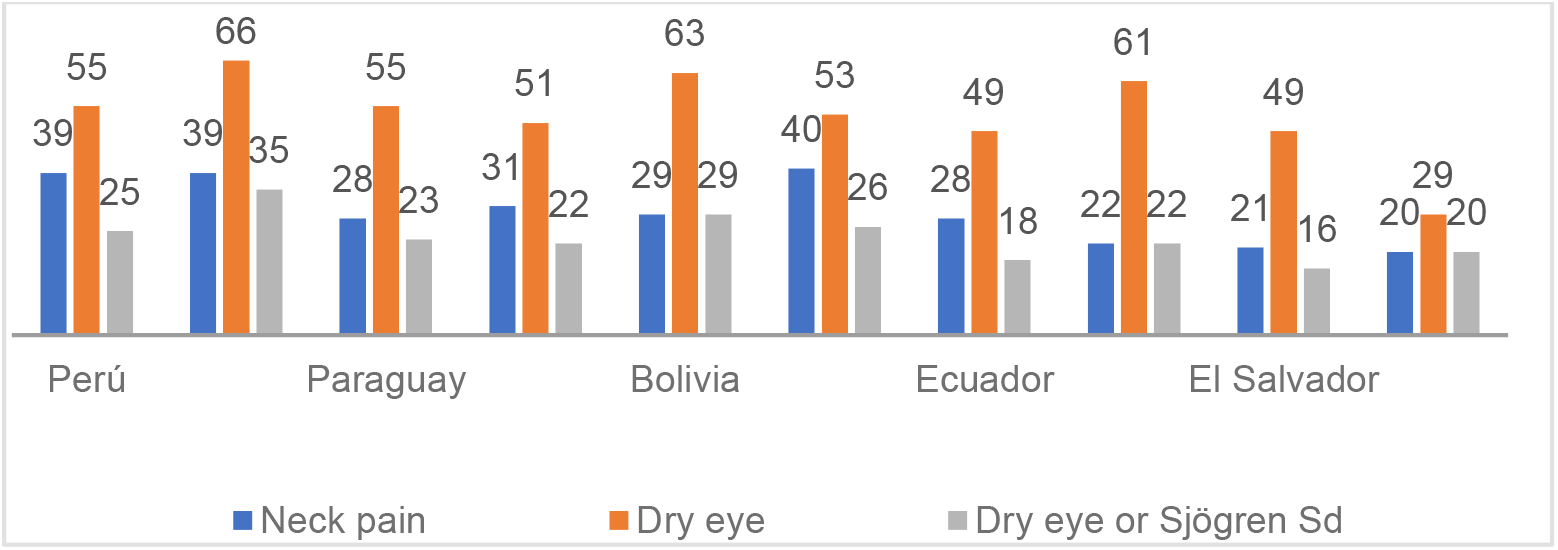
Percentages of neck pain, dry eye and Sjögren’s syndrome by country of residence during the first wave of the pandemic.

When analyzing the factors associated with neck pain, there was more frequency among those with postgraduate studies (RPa: 1,75, IC95%: 1,31-2,33; value p<0,001), among those who were from private universities (RPa: 1,16; IC95%: 1,02-1,33; value p=0,024) and according to the use of more hours a day the computer (RPa: 1,05; IC95%: 1,04-1,05; value p<0,001), Men had less frequency of neck pain (RPa: 0,67; IC95%: 0,59-0,76; value p<0,001), adjusted for year of study, hours of cell phone use and country of residence. **Table 1**

**Tabla 1.**
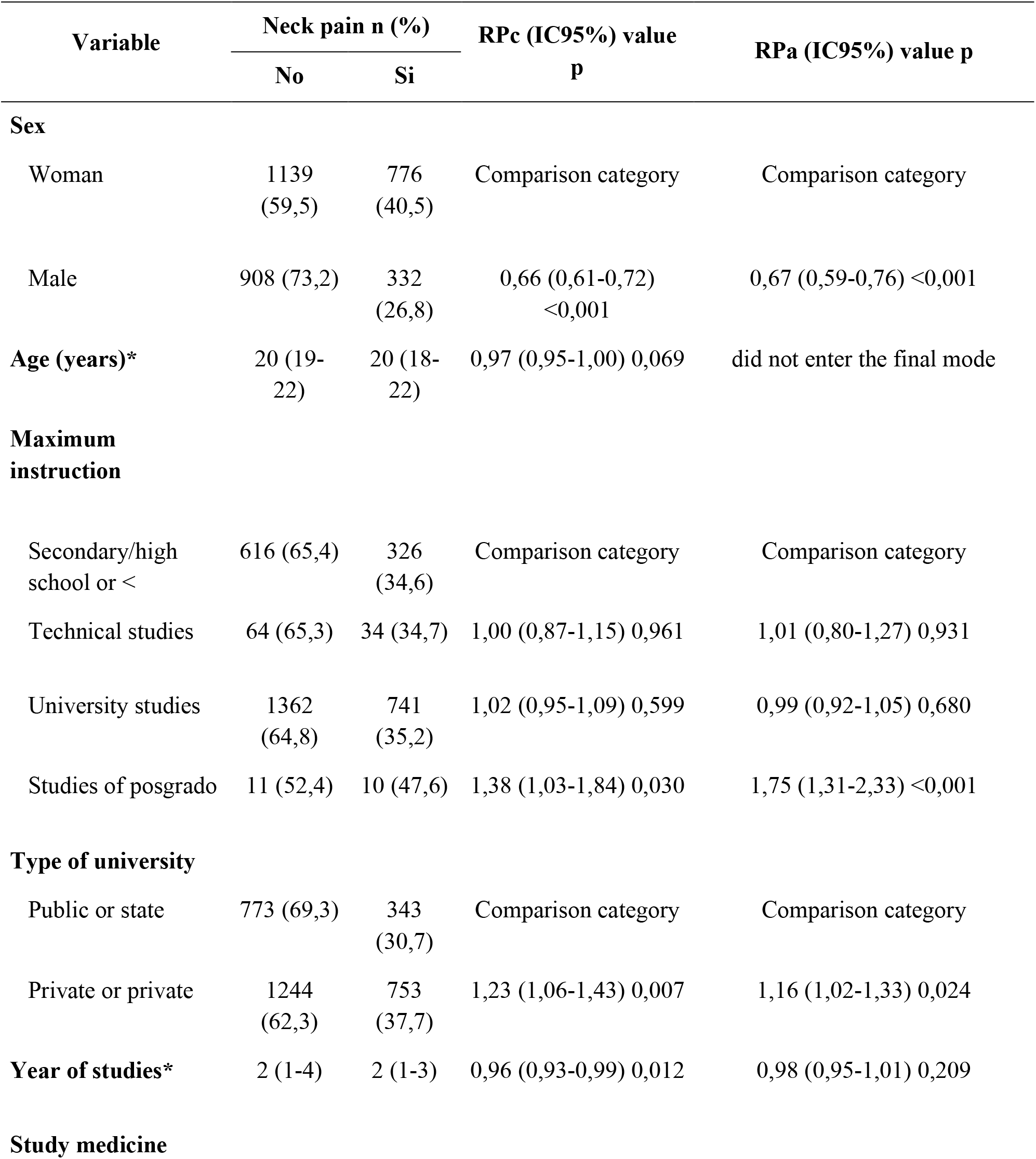
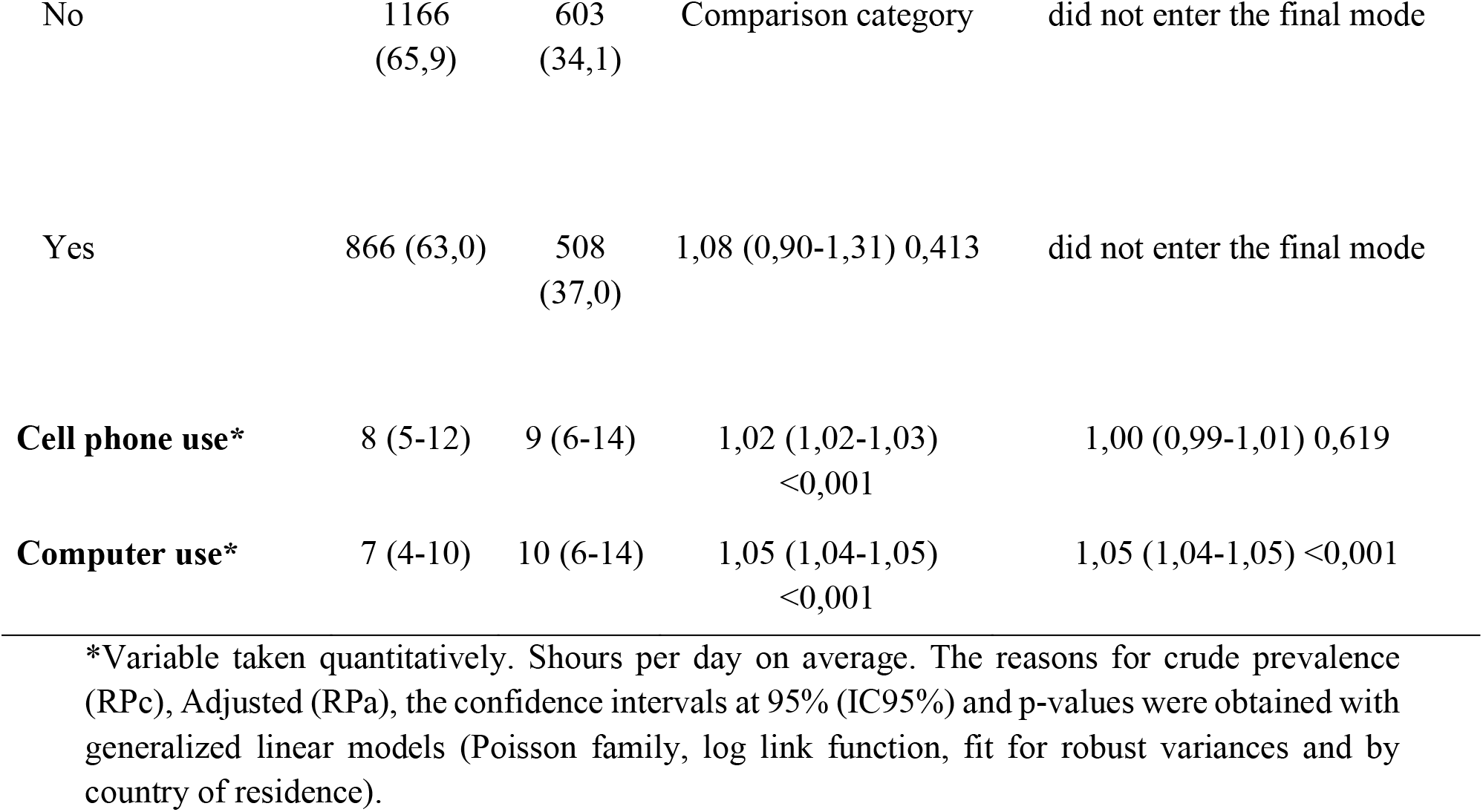
Descriptive and bivariate analysis of factors associated with neck pain among students in Latin America in the first wave of COVID-19.

For factors associated with dry eye, there was more dry eye among those studying medicine (RPa: 1,09, IC95%: 1,01-1,17; valor p=0,030) and according to the use of more hours a day the computer (RPa: 1,02; IC95%: 1,02-1,03; valor p<0,001), In contrast, there was less frequency of dry eye among men (RPa: 0,76; IC95%: 0,69-0,85; valor p<0,001) and university students (RPa: 0,93; IC95%: 0,88-0,99; valor p=0,025); adjusted for age, hours of cell phone use and country of residence. **Table 2**

**Table 2.** Descriptive and bivariate analysis of factors associated with dry eye among students in Latin America in the first wave of COVID-19.

| Variable | Dry eye n (%) |  | RPc (IC95%) Yes p | RPa (IC95%) value p |
| --- | --- | --- | --- | --- |
|  | No | Yes |  |  |
| **Sex** |  |  |  |  |
| Woman | 811 (38,6) | 1291<br>(61,4) | Comparison category | Comparison category |
| Male | 747 (54,2) | 632<br>(45,8) | 0,75 (0,67-0,83)<br><0,001 | 0,76 (0,69-0,85) <0,001 |
| **Age (years)\*** | 20 (19-22) | 20 (19-22) | 0,98 (0,97-0,99) 0,004 | 0,99 (0,97-1,01) 0,466 |
| **Maximum instruction** |  |  |  |  |
| Secondary/high school or < | 446 (42,6) | 600<br>(57,4) | Comparison category | Comparison category |
| Technical studies | 53 (49,5) | 54 (50,5) | 0,88 (0,72-1,06) 0,187 | 0,97 (0,82-1,16) 0,764 |
| University studies | 1048<br>(45,3) | 1267<br>(54,7) | 0,95 (0,92-0,99) 0,011 | 0,93 (0,88-0,99) 0,025 |
| Postgraduate studies | 14 (63,6) | 8 (36,4) | 0,63 (0,38-1,05) 0,075 | 0,67 (0,41-1,07) 0,095 |
| **Type of university** |  |  |  |  |
| Public or state | 594 (47,1) | 668<br>(52,9) | Comparison category | Did not enter the final model |
| Private or private | 947 (43,3) | 1239<br>(56,7) | 1,07 (0,99-1,15) 0,077 | Did not enter the final model |
| **Year of studies\*** | 2 (1-4) | 2 (1-4) | 1,00 (0,97-1,03) 0,769 | Did not enter the final model |
| **Study medicine** |  |  |  |  |
| No | 930 (47,7) | 1019<br>(52,3) | Comparison category | Comparison category |
| Yes | 615 (40,6) | 900<br>(59,4) | 1,14 (1,06-1,22) 0,001 | 1,09 (1,01-1,17) 0,030 |
| **Cell phone use\*** | 8 (5-12) | 8 (5-12) | 1,01 (1,00-1,02) 0,001 | 1,00 (0,99-1,01) 0,603 |
| **Computer use\*** | 8 (4-10) | 8 (5-12) | 1,02 (1,02-1,03)<br><0,001 | 1,02 (1,02-1,03) <0,001 |

\*Variable taken quantitatively. \$hours per day on average. The reasons for crude prevalence (R<sub>Pc</sub>), Adjusted (R<sub>Pa</sub>), the confidence intervals at 95% (IC95%) and p-values were obtained with generalized linear models (Poisson family, log link function, fit for robust variances and by country of residence).

There was a higher frequency of Sjögren’s syndrome among those studying medicine (RPa: 1,22; IC95%: 1,05-1,41; valor p=0,008) and according to the use of more hours a day the computer (RPa: 1,05; IC95%: 1,04-1,07; valor p<0,001), in contrast, men had less Sjögren’s syndrome (RPa: 0,70; IC95%: 0,56-0,87; valor p=0,001); **Table 3**

**Table 3.** Descriptive and bivariate analysis of factors associated with Sjögren’s syndrome among students in Latin America in the first wave of COVID-19.

| Variable | Sjögren's syndrome n (%) |  | R<sub>Pc</sub> (IC95%) value p | R<sub>Pa</sub> (IC95%) value p |
| --- | --- | --- | --- | --- |
|  | No | Yes |  |  |
| Sex |  |  |  |  |
| Woman | 1492 (71,0) | 610 (29,0) | Comparison category | Comparison category |
| Male | 1116 (80,9) | 263 (19,1) | 0,66 (0,55-0,78) <0,001 | 0,70 (0,56-0,87) 0,001 |
| **Age (years)\*** | 20 (19-22) | 20 (18-22) | 0,97 (0,94-1,01) 0,142 | Did not enter the final model |
| **Maximum instruction** |  |  |  |  |
| Secondary/ high school or < | 766 (73,2) | 280 (26,8) | Comparison category | Did not enter the final model |
| Technical studies | 82 (76,6) | 25 (23,4) | 0,87 (0,60-1,26) 0,460 | Did not enter the final model |
| University studies | 1748 (75,5) | 567 (24,5) | 0,91 (0,76-1,09) 0,314 | Did not enter the final model |
| Studies of posgrado | 19 (86,4) | 3 (13,6) | 0,51 (0,22-1,18) 0,116 | Did not enter the final model |
| **Type of university** |  |  |  |  |
| Public or state | 987 (78,2) | 275 (21,8) | Comparison category | Comparison category |
| Private or private | 1594 (72,9) | 592 (27,1) | 1,24 (1,04-1,48) 0,015 | 1,09 (0,93-1,28) 0,286 |
| **Year of studies\*** | 2 (1-4) | 2 (1-4) | 0,97 (0,93-1,02) 0,214 | Did not enter the final model |
| **Study medicine** |  |  |  |  |
| No | 1523 (78,1) | 426 (21,9) | Comparison category | Comparison category |
| Yes | 1066 (70,4) | 449 (29,6) | 1,36 (1,14-1,62) 0,001 | 1,22 (1,05-1,41) 0,008 |
| **Cell phone use\*** | 8 (5-12) | 9 (6-14) | 1,02 (1,01-1,04) 0,002 | 1,00 (0,99-1,02) 0,700 |
| **Computer use\*** | 7 (4-10) | 10 (6-14) | 1,05 (1,04-1,07) <0,001 | 1,05 (1,04-1,07) <0,001 |

\*Variable taken quantitatively. Shours per day on average. The reasons for crude prevalence (RPC), Adjusted (RPa), the confidence intervals at 95% (IC95%) and p-values were obtained with generalized linear models (Poisson family, log link function, fit for robust variances and by country of residence).

## Discussion

The objective was to determine the factors associated with suffering from neck pain, dry eye and Sjögren’s syndrome in Latin American students during the first wave of COVID-19. Many of the Latin American countries had to incorporate online learning into their education system, which caused many alterations in the health of the student population, either at the school level or at the university level.

Our study reported that more hours on the computer were associated with a higher frequency of neck pain, dry eye and Sjögren’s syndrome; students who reported dry eye used the computer on average 8 hours a day, students who had neck pain and Sjögren’s syndrome spent more hours a day in front of the computer (10 hours average and range of 6-14 hours). This is consistent with the research carried out by Espinoza, who evaluated students at the primary level and reported that of the total number of students, 40% spent 3-6 hours and 62% did not use the screen protector; therefore, he concluded that computer use had an impact on the visual acuity of schoolchildren. (22). Another study mentions that even before receiving a virtual education, there were already pathologies / ailments that were related to prolonged time in front of a computer (13).

Neck pain was more prevalent among women (13% more compared to men), as well as a higher prevalence of dry eye (15% more compared to men) and Sjögren’s syndrome (10% more compared to men), in a study among university students on head-neck pain and the degree of disability in relation to computer use, It was reported that the female population was the most affected, in addition, the statistical analysis showed a high prevalence (57% with neck pain and 43% with neck pain plus headache); having a relationship according to the number of hours that the computer was used and the practice of sport, however, that no relationship was found between headache and computer use (23). Female gender also had higher risk in Asian study (p= 0,026), stating that women had more negative effects (24). This could be because hormonal changes occur in women during the reproductive cycle and many students use hormonal contraceptives, which could cause a variation in the lipids and proteins of the meibomian glands, which would condition dry eye (25). This needs further research, because if so, women should be instructed to protect themselves more carefully, which could lead to significant long-term pathologies.

Medical students had more frequent dry eye and Sjögren’s syndrome. A fact that was also demonstrated in a Peruvian study, where male medicine students between the ages of 16-23 years had worse visual health, who spent more than 6 hours a day in front of the computer and more than 5 hours in front of the cell phone (26). In Thailand a study was associated with studying medicine or to be paramedics with Dry Eye Syndrome, which could be due to the great psychological stress experienced in the pandemic (24), In addition, students diagnosed with Sjögren’s syndrome use the computer between 4-10 hours a day, as well as the cell phone between 6-14 hours; so, we can say that the greater the number of hours on the cell phone and computer there is a greater probability of suffering from Sjögren’s syndrome. It is known that the coronavirus pandemic led people to opt for teleworking, as well as online education, allowing workers and students to continue their work remotely; however, excessive use of some devices can lead to visual problems (27). It is recommended to evaluate other careers, to expand this work, since there was an important access to medical students, this because the main authors are doctors, however, this limitation was partial, because there was also a significant number of respondents from other careers, which would be a starting point for other researchers to measure these realities in other areas, professions, or occupations.

As for the degree of instruction, graduate students had more neck pain, but high school students had a higher frequency of dry eye, there may be different causes that condition this pathology and affect a certain group, especially this may be related to the longer time of use of the computer or cell phone, as well as, bad postures in front of these devices (27, 28). High school students compared to other students of different grade/instruction reported having dry eye symptoms in 45%, this unlike university students, who presented more dry eye or Sjögren’s syndrome; as reported in another study, where high school students reported headaches, neck pain and also visual problems (29). This is related to what was reported in Saudi Arabia, where 50% of the participants of students were not able to have control over their emotions, overvaluing their link with cell phones, using them even before doing any other activity, which increased their possibility of developing Dry Eye Syndrome and also generates a distortion of body mechanics; causing muscle aches in the neck, shoulders, back and whole body (30).

In private universities a higher percentage of neck pain was found, although this difference was only 7% with respect to those of public universities, this was statistically significant when adjusted with other variables in the final model. There are still not many results of this type, which find differences between the presentation of neck pain according to whether they belonged to a public or private school, but if it has been reported that there are differences between workers of private companies according to their time of exposure to screens, so more research should be done according to the type of context in which students and even professionals are, since, it is known that institutions vary in their way of providing protection mechanisms, training and even the motivation to protect themselves (12, 13). There are descriptive reports that show this reality in only one type of population, for example, in a private Peruvian university musculoskeletal pain and postural load due to work were evidenced; being the most affected area located at the cervical level (79%); with a pain time greater than 7 days, inadequate postures in 66% of respondents and 45% with a high risk level (31). These differences may have been generated by the conditions offered by private universities, since, according to a study, it showed that these institutions had good coverage and access to virtual education, in addition, to perceive that their teachers were more qualified in front of information technologies (32), which would facilitate the adequate teaching of virtual classes and, therefore, with less repercussions than those of public universities. Future research is expected to help find more answers among these groups of respondents.

The results found are very interesting, since they show a reality six months after the beginning of the most disastrous stage for many Latin American countries during the pandemic, although there are limitations that the results cannot be extrapolated to all the students, countries or study centers from which the data came (this because that objective was not met, but if the power to find important associations), many of the respondents were from medicine (or health sciences) and could not have many other variables that could influence the presentation of these pathologies (such as the use of ergonomic elements, if their university gave them training, the way of using the elements, among others). Despite all the limitations, it was possible to survey more than three thousand students throughout Latin America, and the vast majority of crossings had a very good power, so these results can give new policies for the management of this problem, which post pandemic would continue to be generated, since, wearer facing A new reality, especially because many schools have incorporated virtuality as part of teaching. For this reason, it is recommended that more studies be carried out on the subject, in specific populations, with diverse interventions and with varied methodological designs, so that new forms of intervention are available to minimize this problem among the student community and other populations that present it.

## Conclusion

For all the above, it is concluded that the three pathologies / ailments were associated with the greatest number of hours of computer use and sex, medical students had more frequencies of dry eye and Sjögren’s syndrome, graduate students had more neck pain, but university students had less dry eye and those from private universities had more neck pain.

## Data Availability

All data produced in the present study are available upon reasonable request to the authors

## Author Contributions

Conceptualization CRM, BGL, ILZL, DAC, MFF, CAV, MAVE, VSA, TR, LIZ, EET. Methodology C.R.M validation C.R.M, formal analysis, C.R.M, data curation: C.R.M, writing original draft preparation: CRM, BGL, ILZL, DAC, MFF, CAV, MAVE, VSA, TR, LIZ and EET, writing-review and editing: CRM, BGL, ILZL, DAC, MFF, CAV, MAVE, VSA, TR, LIZ and EET, visualization: CRM, BGL, ILZL, DAC, MFF, CAV, MAVE, VSA, TR, LIZ and EET

## Institutional Review Board Statement

The study was conducted under the Declaration of Helsinki. This research’s preparation and execution fully complied with the fundamental ethical principles of autonomy, justice, beneficence, and non-maleficence. The Act Number (0013-2022-UPAO), approved by the Bioethics Committee of the Universidad Privada Antenor Orrego.

## Data Availability Statement

The data presented in this study are available on request from the corresponding author.

## Acknowledgments

To the surveyors of the work, who supported very actively during the execution of the research, as well as to the Norbert Wiener Private University, which financed the process of quality control of the data.

